# How many COVID-19 cases could have been prevented in the US if its interventions were as effective as those in China and South Korea?

**DOI:** 10.1101/2020.05.06.20092981

**Authors:** Kai Liu, Yukun Song, Menghui Li, Zhesi Shen, Ming Wang, Jinshan Wu

**Author notes:** Corresponding author *Email address* (Jinshan Wu).

## Abstract

The COVID-19 [1] pandemic has forced governments to take measures to contain the spread of the disease [2]; however, the effects have varied significantly from one country to another contingent on governments’ responses. Countries that have flattened their coronavirus curves prove that interventions can bring COVID-19 under control. These achievements hold lessons, such as the strict social distancing and coordinated efforts of all government levels in China and massive testing in South Korea, for other countries battling the coronavirus around the world. In this work, we attempt to estimate how many COVID-19 cases could have been prevented in the United States (US) when compared with the US’s actual number of cases assuming that on a certain date, the US took China-like or South Korea-like interventions and that these interventions would have been as effective in the US as in China and South Korea. We found that if that date was at the early stage of the outbreak (March 10), more than 99% (1.15 million) fewer infected cases could be expected by the end of the epidemic. This number decreases to 66.03% and 73.06% fewer infected cases with the China-like scenario and the South Korea-like scenario, respectively, if actions were taken on April 1, highlighting the need to respond quickly and effectively to fight the virus. Furthermore, we found that although interventions in both China and South Korea allowed the COVID-19 outbreak to be managed, the epidemic could still oscillate without strict large-scale ‘lockdown’ measures, as shown in South Korea. Our results demonstrate that early effective interventions can save considerably more people from infection and provide a worldwide alert regard the need for swift response.

---

The outbreak of COVID-19 is spreading in most regions around the world and has been declared a global pandemic [3]. According to the World Health Organization (WHO), as of 3 April 2020, the world approached the one million mark of reported cases of COVID-19 with a death toll of over 60 thousand [4]. While some countries are experiencing exponential growth in case numbers, several countries, such as China and South Korea, have successfully controlled the spread of COVID-19. As stated by Dr. Tedros of the WHO, ‘The experience of these countries and of China continues to demonstrate that this is not a one-way street. This epidemic can be pushed back, but only with a collective, coordinated and comprehensive approach that engages the entire machinery of government’[5]. Considering the advanced biotechnology and ICU capacity per citizen (20.0–31.7 ICU beds per 100,000 inhabitant [6], higher than any other country in the world) in the US, the large case number (431, 437 as of April 8) and the high death rate (3.4% as of April 8) of COVID-19 in the US seem puzzling [7]. The fact that the United States and South Korea announced their first cases of COVID-19 on the same day [7] makes the comparison between the US and South Korea even more valuable. As noted by Dr. Anthony Fauci [8], the U.S. government’s top infectious disease expert, the US “could have saved lives” if it had introduced measures to stop COVID-19 earlier. In this work, we aim to answer the question of the extent to which infections could have been reduced if interventions in the US had been introduced earlier and were as effective as those in China and South Korea.

We perform this estimation by making use of the empirical values of the time-varying reproduction number (*R*(*t*)), which is the average number of cases infected by one infector at time [9, 10] of the US *(R_us_*(*t*)), China *(R_cn_*(*t*)) and South Korea (*R_kr_*(*t*)). We consider the latte *R_us_(t)* two typical countries where the spread of COVID-19 has already been controlled. *R* (*t*) > 1 indicates that the outbreak is self-sustaining, while *R*(*t*) < 1 indicates that the number of new cases decreases over time and will eventually end [11]. At a certain date *τ*, when we assume that the US takes measures that are as effective of those in one of the two typical countries, such as China, we connect *R_us_*(*t* ≤ *τ* − 1) and *R_cn_*(*t*) to form *R_us–cn_* (*t*; *τ*) for *t* > *τ*, and the accumulated number of cases *C_us−cn_* (*t*; *τ*) at time t is calculated based on *R_us–cn_ (t; τ*) (see Methods). By comparing the actual accumulated case numbers at time t in the US, *C_us_ (t > τ*) and *C_us–cn_ (t > τ; τ*), we estimate how many cases could have been prevented at time t in the US if actions were taken at time τ and if the actions were as effective as those in typical countries.

We first show the time-varying reproduction number of different countries, *R_cn_*(*t*), *R_kr_(t)* and *R_us_*(*t*). As we can see from Fig. 1(a), the time-varying reproduction numbers in China *R_cn_*(*t*) showed a very rapid decreasing starting *R* = 2.63 (95%CI: 1.96 − 3.39) in the early stages of the epidemic to *R* = 0.98 on February 6, which continued to decrease and subsequently remained at a very small value. The effective reproduction number of South Korea, *R_kr_*(*t*), was 3.34 (95%CI: 3.13 − 3.56) initially (Fig 1(b)) and declined to less than 1.0 on March 8. The value of *R* bounced back to larger than 1.0 on March 31 and stayed near that level from that time.

**Figure 1:**
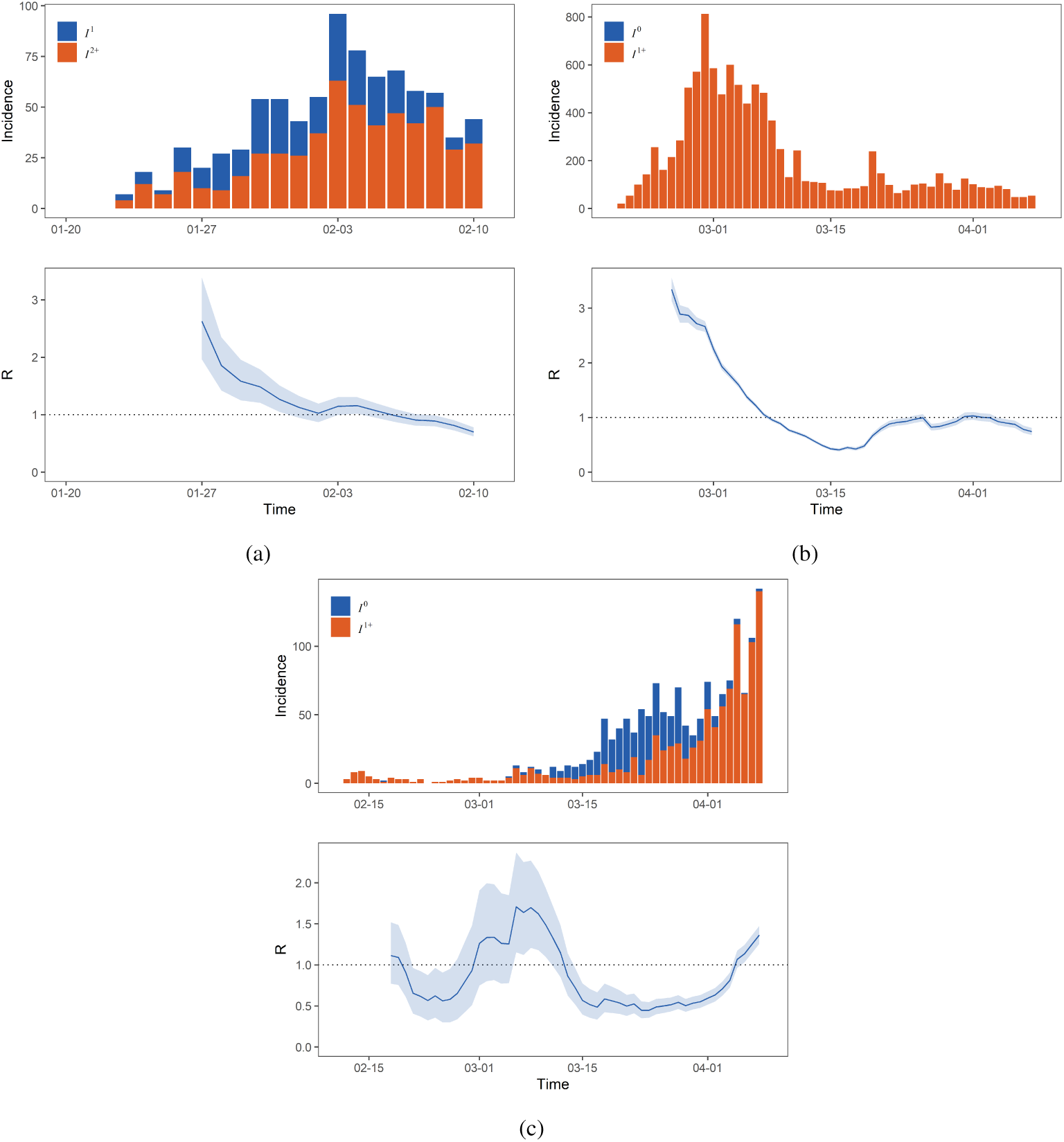
Evolution of the number of cases and the time-varying reproduction number *R*(*t*) in China (a), South Korea (b), and Singapore (c). *I*^0^ (*t*), *I*^1^ (*t*) and *I*^2^ (*t*) denotes respectively the imported thus the zeroth-generation cases, the local cases infected by the imported cases thus the first-generation cases, and the cases infected by the first-generation cases, and the cases infected by the first-generation cases thus the second-generation cases. A notation *I^n^*^+^ (*t*) refers to the *n*-th and higher generations. Shaded regions of *R*(*t*) mark the 95% prediction envelops.

The patterns of *R*(*t*) in China and South Korea are closely related to its implemented interventions. As the country that first reported the virus, China has taken the most comprehensive, strictest and most thorough measures to battle the epidemic [12]. These measures include the Wuhan lockdown, isolating suspected and confirmed cases, closing schools and entertainment venues, quarantine of residential communities or villages, the declaration of travel experiences and self-isolation at home for two weeks [13, 14, 15, 16, 17]. China has done so with the coordinated efforts of all government levels and at the level of individuals. The 1.4 billion Chinese people, regardless of whether they are infected, suspected or not, understand and cooperate with the strict interventions to stop the spread of the virus.

South Korea has been able to control the epidemic without massive quarantine and with relatively less disruption to daily living [18, 19]. The outbreak around February 19, partially due to religious groups in South Korea, was quite serious [20] perform the most massive testing of the virus in the world together with transparent case reporting and extensive contact tracing efforts [21], which led to a dramatic decrease of *R*.

Although both the Chinese and South Korean approaches have enabled the COVID-19 outbreak to be managed to date, the patterns of *R* suggest that without large-scale “lockdown” measures as in China, *R* might still oscillate, and it is difficult to maintain *R* < 1, as shown in South Korea. Singapore, which took a similar approach to South Korea, is a good illustration. It responded swiftly to its first case of coronavirus on January 23 [22] and implemented measures such as limiting airlines, large-scale testing, and strict isolation policies [23, 24, 25]. The time-varying reproduction numbers in Singapore *R_sg_* (*t*) fluctuated with a mean value of 1.0 from February 18-March 12 due to its quick and efficient response and peaked with a value of 1.71(95%CI: 1.15 − 2.37) on March 6 (Fig. 1 c). This value declined to below 1. 0 after March 12 but gradually climbed to more than 1.0 from April 5, leading to a second outbreak of infections.

Fig. 2(a) shows the evolution of the number of cases and the time-varying reproduction number *R_us_* (*t*) of the US. Without enforcing social distancing measures and other mitigation strategies [26], the *R* of the US reached as high as 4.16(95%CI: 3.83 − 4.51) for the early phase with large fluctuations.

**Figure 2:**
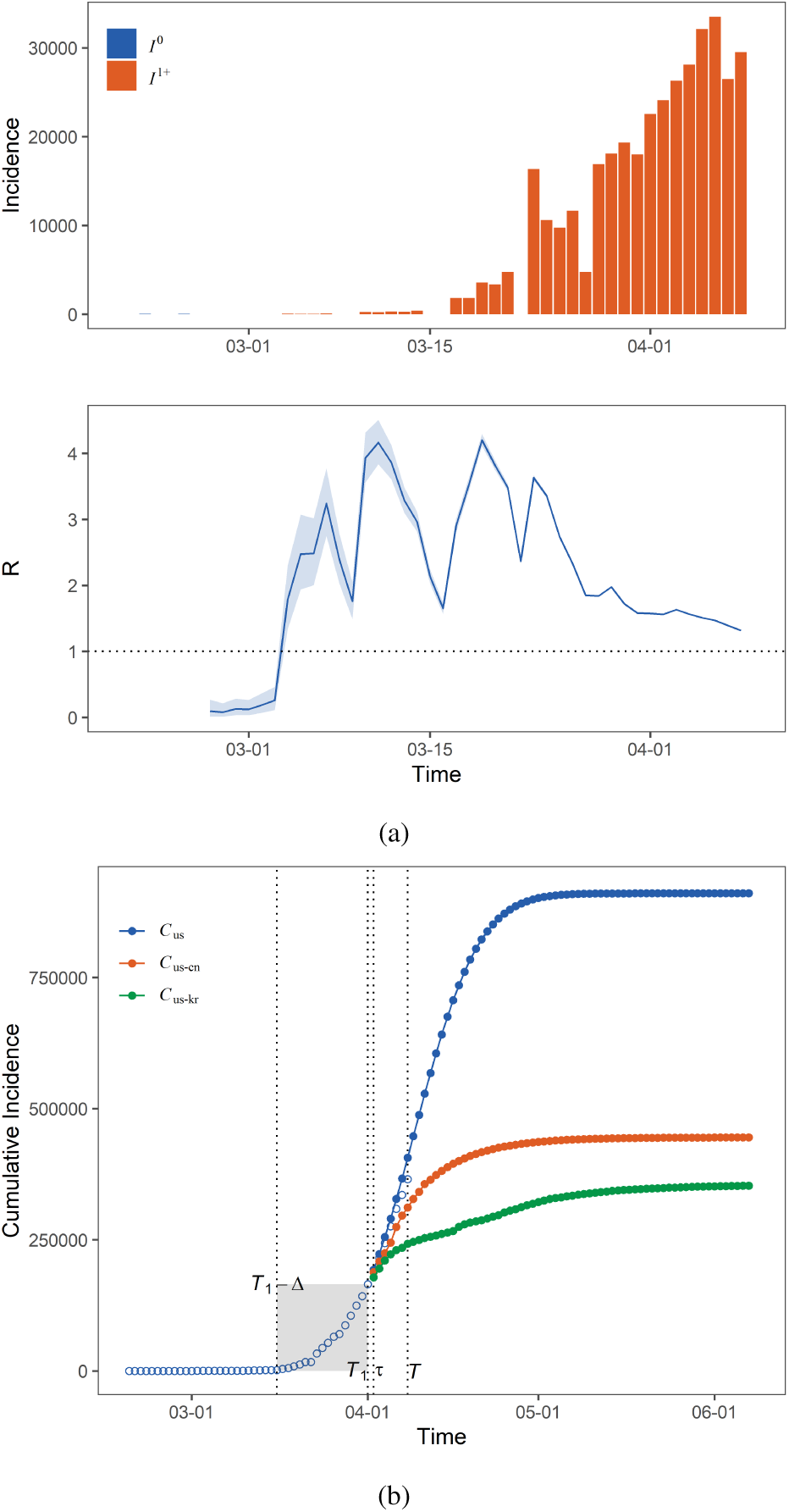
(a) Evolution of the number of cases and the time-varying reproduction number *R(t)* in the US. Shaded regions of *R* (*t*) mark the 95% prediction envelops. (b) Accumulated case numbers at a time *T* and at the end of epidemics based on extrapolated *R_us_* (*t*) (blue line), *R_us–cn_* (*t, τ*) (orange line), and *R_us–kr_ (t, τ)* (green line). *τ* indicates the time that effective actions were taken. The extrapolated *R_us_* (*t*) makes use of actual data in *t* ∈ [*T*_1_ − ∆, *T*_1_], where *T*_1_ < *T* so that we can compare the extrapolated *R_us_* (*t*) and the actual *t* ∈ [*T*_1_ − ∆, *T*_1_].

With these *R_cn_* (*t*) and *R_kr_* (*t*), we then form *R_us−cn_* (*t, τ*) (*R_us−kr_* (*t*, *τ*)) and calculate the accumulated case numbers *C_us−cn_* (*t* = *T*, *τ*)(*C_us−kr_* (*t* = *T*, *τ*)) at a time *T* and at the end of epidemics *C_us–cn_* (*t* = ∞, *τ*)(*C_us−kr_* (*t* = ∞, *τ*)). *τ* indicates the time that the US took a China-like or South Korea-like approach and those measures were as effective as in these two countries. We also extrapolate *R_us_* (*t*) to obtain an estimate of *C_us_* (*t* = ∞) from the already known *R_us_* (*t* ≤ *T*) to *t* ≫ *T*. Details of this extrapolation are reported in the Methods section. For *t* ≤ *T*, we have *C_us_* (*t*), the actual number of cases in the US.

In Fig. 2 (b), we show that if *τ* =April 2, on April 8, the daily case number would have been near 311, 657 and 242, 488, respectively, 14.6% and 33.6% lower than the actual numbers of cumulative cases, which is the actual number on April 8, in the US. We can also compare the accumulated case numbers at the end of the epidemics until *R* = 0. Fig. 2 (b) shows the difference between *C_us_* (*t* = ∞) = 909, 976, *C_us–cn_* (*t* = ∞; *τ*) = 445, 089 and *C_us–kr_* (*t* = ∞; *τ*) = 352, 980.

We further estimated how many infections could have been avoided if the US had taken effective actions at earlier stages of the spread. In Fig. 3, we report the difference between the accumulated case numbers, for example, *C_us–cn_* (*t* = *T*; *τ*) and *C_us_* (*t* = *T*) as a function of *τ* and *C_us–cn_* (*t* = ∞; *τ*) and *C_us_* (*t* = ∞) at the end of the epidemics. Clearly, the number of infections could be reduced with early interventions. We can see that if effective actions such as those in China were taken at *τ* = March 10, which was two weeks after the first coronavirus case that did not have known ties to an existing outbreak in the US [27], at the current day *T* =April 8, a total of 364,602 (99.86%) infected cases could have been reduced, and by the end of this epidemic, a cumulative number of 1,155, 620 ± 181, 006 (99.96%) infected cases could be reduced. Similarly, if South Korea-like actions were taken on the same date *τ* =March 10, then 364, 720 (99.89%) cases at time *T* =April 8 and 1,155, 713 ± 181,007 (99.96%) cases at the end could be avoided. Table 1 shows the number of potentially reduced cases for several values of τ. The avoidance impact would be reduced if actions were not taken in a timely fashion; only 66.03% and 73.06% fewer infected cases would be achieved with the China model and the South Korea model, respectively, when *τ* =April 1.

**Figure 3:**
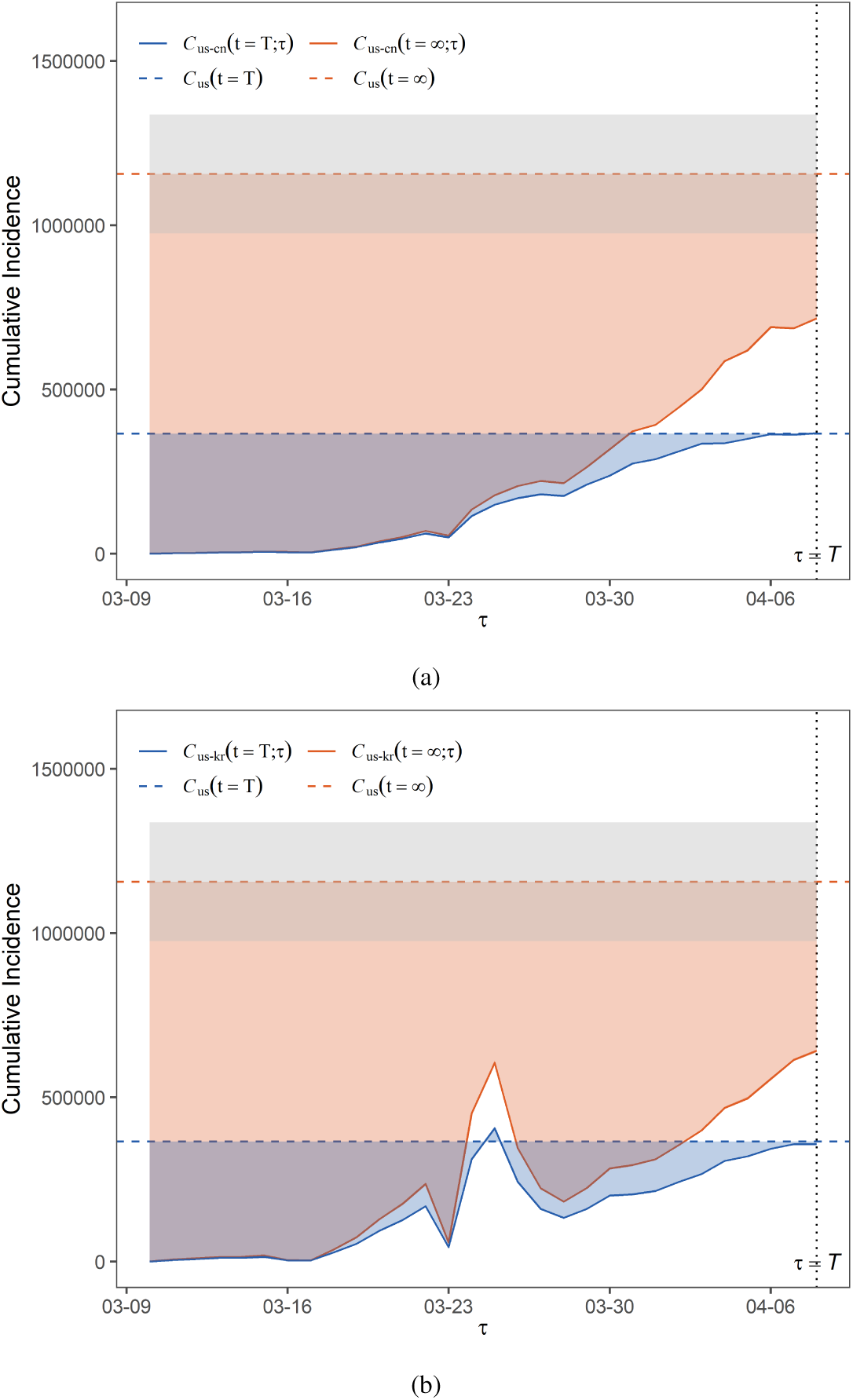
Evolution of the reduced infections in the US with *τ* based on *R_us−cn_* (*t*, *τ*) (a) and *R_us_*_−_*_kr_ (t, τ)* (b). Blue and orange dotted lines indicate *C_us_ (T)* the actual infected number in the US at *T* = April 8 and *C_us_* (∞) the estimated infected number at the end of epidemic based on the extrapolated *R_us_* (*t*). We also plot around *C_us_* (∞) with gray regions mark one standard deviation (see the Method section for details). Blue and orange shaded region indicate the reduced infected number at *T* = April 8 and at the end of epidemic, respectively, if effective actions were taken at time *τ*.

**Table 1:**
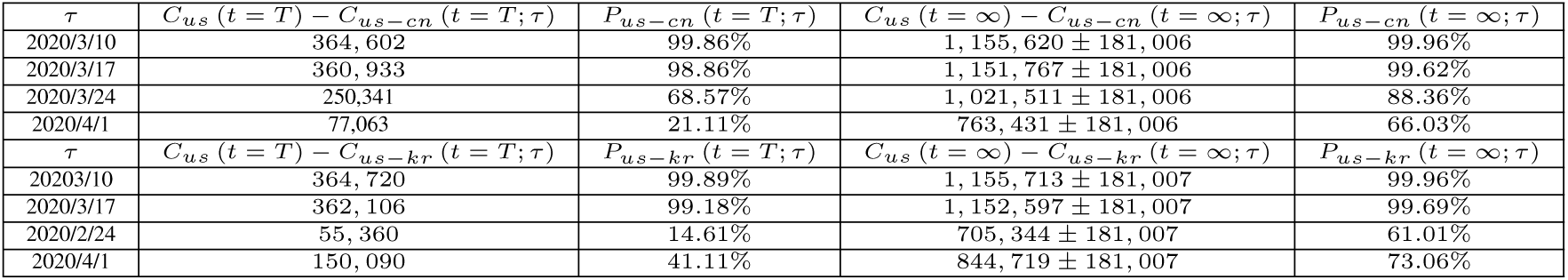
Reduced infections in the US if effective actions were taken at several time spots τ: reduced infected cases *C_us_* (*t* = *T*) − *C_us−cn_* (*t* = *T*; *τ*)(*C_us_* (*t* = *T*) − *C_us−kr_* (*t* = *T*; *τ*)) and reduced percentage of infected cases *P_us−cn_ (t = T; τ) (P_us−kr_ (t = T*; *τ*)) at time *T* =April 8, reduced infected cases *C_us_*(*t* = ∞) − *C_us−cn_* (*t* = ∞; *τ*) (*C_us_* (*t* = ∞) − *C_us−kr_* (*t* = ∞; *τ*)) and reduced percentage of infected cases *P_us−cn_* (*t* = ∞; *τ*) (*P_us−kr_* (*t* = ∞; *τ*)) at the end.

| $\tau$ | $C_{us}(t=T) - C_{us-cn}(t=T; \tau)$ | $P_{us-cn}(t=T; \tau)$ | $C_{us}(t=\infty) - C_{us-cn}(t=\infty; \tau)$ | $P_{us-cn}(t=\infty; \tau)$ |
| --- | --- | --- | --- | --- |
| 2020/3/10 | 364,602 | 99.86% | $1,155,620 \pm 181,006$ | 99.96% |
| 2020/3/17 | 360,933 | 98.86% | $1,151,767 \pm 181,006$ | 99.62% |
| 2020/3/24 | 250,341 | 68.57% | $1,021,511 \pm 181,006$ | 88.36% |
| 2020/4/1 | 77,063 | 21.11% | $763,431 \pm 181,006$ | 66.03% |
| $\tau$ | $C_{us}(t=T) - C_{us-kr}(t=T; \tau)$ | $P_{us-kr}(t=T; \tau)$ | $C_{us}(t=\infty) - C_{us-kr}(t=\infty; \tau)$ | $P_{us-kr}(t=\infty; \tau)$ |
| 2020/3/10 | 364,720 | 99.89% | $1,155,713 \pm 181,007$ | 99.96% |
| 2020/3/17 | 362,106 | 99.18% | $1,152,597 \pm 181,007$ | 99.69% |
| 2020/2/24 | 55,360 | 14.61% | $705,344 \pm 181,007$ | 61.01% |
| 2020/4/1 | 150,090 | 41.11% | $844,719 \pm 181,007$ | 73.06% |

Our analysis shows the effects of interventions implemented in China and South Korea based on an investigation of the time-varying reproduction number in each of the countries. The extremely strict approach that China employed allowed the reproduction number to be stably below 1.0; this approach contained the virus spread in China and prevented it from spreading further. The aggressive massive testing of viruses together with other less strict measures also reduced the reproduction numbers with possible fluctuations. If similar approaches had been adopted in the US at the early stage of outbreak, e.g., two weeks after the first case without clear ties to an existing outbreak, more than 99% fewer infected cases (1.15 million) could be expected for the US by the end of the epidemic under both the China model and the South Korea model.

The basic idea of our analysis, that is, to connect the time series of the reproduction numbers of China and the US, has its own limitations. In principle, one should develop a model with the existing conditions [28, 29] of the US and modify the parameters in the US model by conceptualizing the key strategies in China or South Korea to determine how many infected cases there will be at the end or at a time in the middle. However, developing such a model requires many parameters and unknown information, which may be time-dependent and have high uncertainty. Therefore, in this work, we simply assume that regardless of the strategies, they will have the same effectiveness in China or South Korea as in the US and connect the time series of the reproduction numbers *R_us_* (*t*) to *R_cn_*(*t*) or *R_kr_* (*t*). This is an oversimplification. Another potential issue is the extrapolation of the reproduction numbers *R_us_*(*t*) for *t* > *T*, where T is the last date of our data. Again, in principle, one should also develop an epidemic model for that prediction. However, since we only want to obtain a reference value for the worst-case scenario, which is used to compare with better-case scenarios directly connecting *R_us_* (*t*) to *R_cn_* (*t*) or *R_kr_* (*t*), and because the foreseen results from this simple extrapolation are acceptable in a short time window, we choose to use this rather simple approach.

We do not aim to investigate whether the measures of China and South Korea can be replicated in the rest of the world or to compare which interventions are most effective. Rather, we offer perspective for other countries, using the US as an example in this paper, to identify what could occur if approaches similar to those in the above typical countries were taken. Results of other countries in addition to the US can be found in the supplementary file. In fact, each country has its own unique challenges in the face of the outbreak, and there is no successful experience in one country that can be completely reproduced in another country. Other countries need to consider their own situation, including outbreak severity, testing regimes, medical capacities, the health-care system, and culture, before deciding on a direction. However, every country must act quickly and powerfully to fight the virus, and any government’s inaction with regard to epidemics could pose a serious challenge to global public health. We have seen that some countries missed a window of opportunity to control coronavirus, but it is never too late to take actions. Policy decisions made today will determine the extent to which the spreading effects of virus and timely and effective interventions could help to end the outbreak early.

## Data Availability

A report on the raw data and a way of sharing those cases with generation labels will be published elsewhere; prior to their publication, a very rough version can be obtained at GitHub(https://github.com/Bigger-Physics/COVID19-reducedcases) or via email request.

## Acknowledgements

This work is jointly supported by the National Natural Science Foundation of China under grant nos. 41330423 and 41420104006.

## Author contributions

Jinshan Wu and Kai Liu designed the research. Yukun Song, Menghui Li, and Zhesi Shen performed the analysis. Kai Liu formed the draft with the contribution from Jinshan Wu. Jinshan Wu and Ming Wang revised the draft. All authors contributed to the interpretation of the results and the improvement of the paper. Special thanks to Jiatong Zhu, Qianzhi Wang, Jianxin Zhang, Kaiwen Li, Yuting Zhang, and Ningning Qiao for collecting and clearing the data.

## Methods

### Data

Our data on US cases are obtained from the WHO, and we obtain our data on Chinese cases by extracting information from the official reports released by provincial/municipal health commissions in China. The reason we cannot directly use the WHO data on Chinese cases is that, first, we need the reproduction number of transmissions of local cases instead of imported cases, which is clearly different in China; second, the data quality of early Hubei cases is not as good as that in other provinces. From the official reports, we classify all cases when possible into imported cases, first-generation cases that are infected by the imported cases, and second-generation and beyond cases, which are infected by the first-generation and beyond cases. Of course, by doing so, we discarded a portion of cases for which we could not determine the generation number. To reduce the bias of this step on the reproduction number, we only make use of the data in those provinces where a large portion of the cases have been classified into generations. Ultimately, we arrived at a dataset of Chinese cases in 11 provinces where, on average, 85% cases have been identified. A report on the raw data and a way of sharing those cases with generation labels will be published elsewhere; prior to their publication, a very rough version can be obtained at GitHub(https://github.com/Bigger-Physics/COVID19-reducedcases) or via email request.

**Equations between** *I (t)* **and** *R* (*t*). We use EpiEstim 2 [30] and the formula behind it for this calculation. Given a time series of imported cases denoted as *I*^0^ (*t*), which can be zero if there are no imported cases for this region under consideration, and a time series of local cases denoted as *I*^1^ (*t*), the time-varying reproduction number at time t, denoted as *R* (*t*), can be calculated from

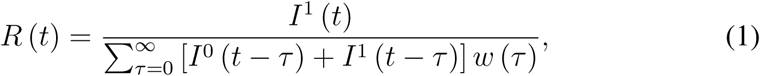

where *w(τ*) is the generation interval (GI) distribution of the epidemics, which indicates the likelihood that an infector will infect a new infectee after *τ* time after the infector is infected. When GI is not available, serial interval (SI) is often used instead, although for epidemics with per-symptomatic transmissions, such a replacement is questionable [31]. However, in this work, we make use of SI instead of GI, and the SI data are obtained from ref [32]. With this known SI, given the full time series *I*^0^ (*t*) and *I*^1^ (*t*), we can find *R* (*t*) using Eq. 1.

Correspondingly,

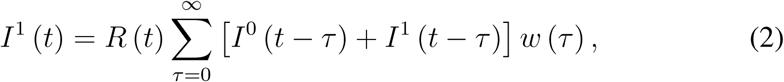

and the total number of daily cases includes both the local and imported cases,

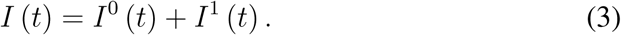

With this known SI, given the full time series *R* (*t*) and *I*^0^ (*t*) together with historical data of *I*^1^ (*τ* < *t*), we use Eq. 3 to calculate the number of daily cases including both the local and imported cases. In our extrapolation, when there are no data on the future daily imported cases, we focus only on local cases; thus, we use Eq. 2.

Estimating *R_cn_* (*t*) is trickier since due to the different interventions, the effective reproduction number of imported cases and local cases in China is clearly different. Therefore, we have extended the framework for estimating the reproduction numbers; that extension will be reported elsewhere. For now, we present the final formula:

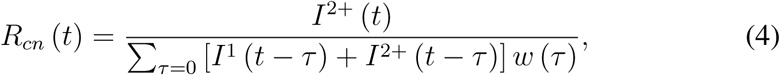

where *I*^2+^ (*t*) is the number of cases with second-generation and beyond that are infected by the first-generation and beyond local cases.

Once we have the daily case number *I* (*t*), we can use the accumulated case number and vice versa:

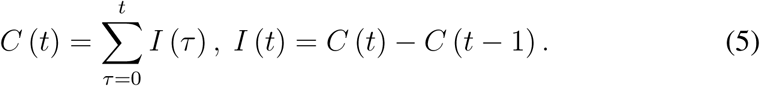

**Extrapolating** *R_us_* (*t*). Sometimes we need to extrapolate *R_us_* (*t* ≤ *T*) to *R_us_* (*τ*) at a time *τ* > *T*. The daily values of *R_us_* (*t*) are oscillating, which makes extrapolation very difficult. We then apply a *W–*day (*W* = 7 through this work) time window average of the daily *R_us_* (*t*) to obtain an 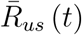. We then take the 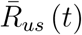 for *t* ∈ (*T* − ∆, *T*] in a window with size ∆ and use it for extrapolation. In choosing this Δ, we attempt to include the longest possible period of time before time *T* but still with a more or less monotonically increasing or decreasing trend. We then fit this value of 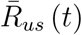 with the following decreasing function 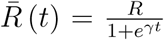, where the parameters *R*, *γ* will be obtained from the fitting. Then, the fitted curve is used to generate future values of 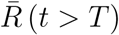 and let 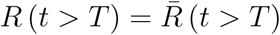.

As mentioned previously, in principle, this fitting may be very far from the actual pattern of the time-varying reproduction number of the US. However, when we take a short time window so that within the window 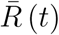 is monotonic, we can see that the extrapolated results are not that bad. In Fig. 2(b) of the main text (and also Fig.4 (b), Fig.6(b) and Fig.8(b)), we provide examples of the extrapolated results for *t* ∈ (*T*_1_ − ∆, *T*_1_] for *T*_1_ < *T* so that we have empirical data between *T*_1_ and *T* to determine how far the extrapolated results are from the actual data.

**Figure 4:**
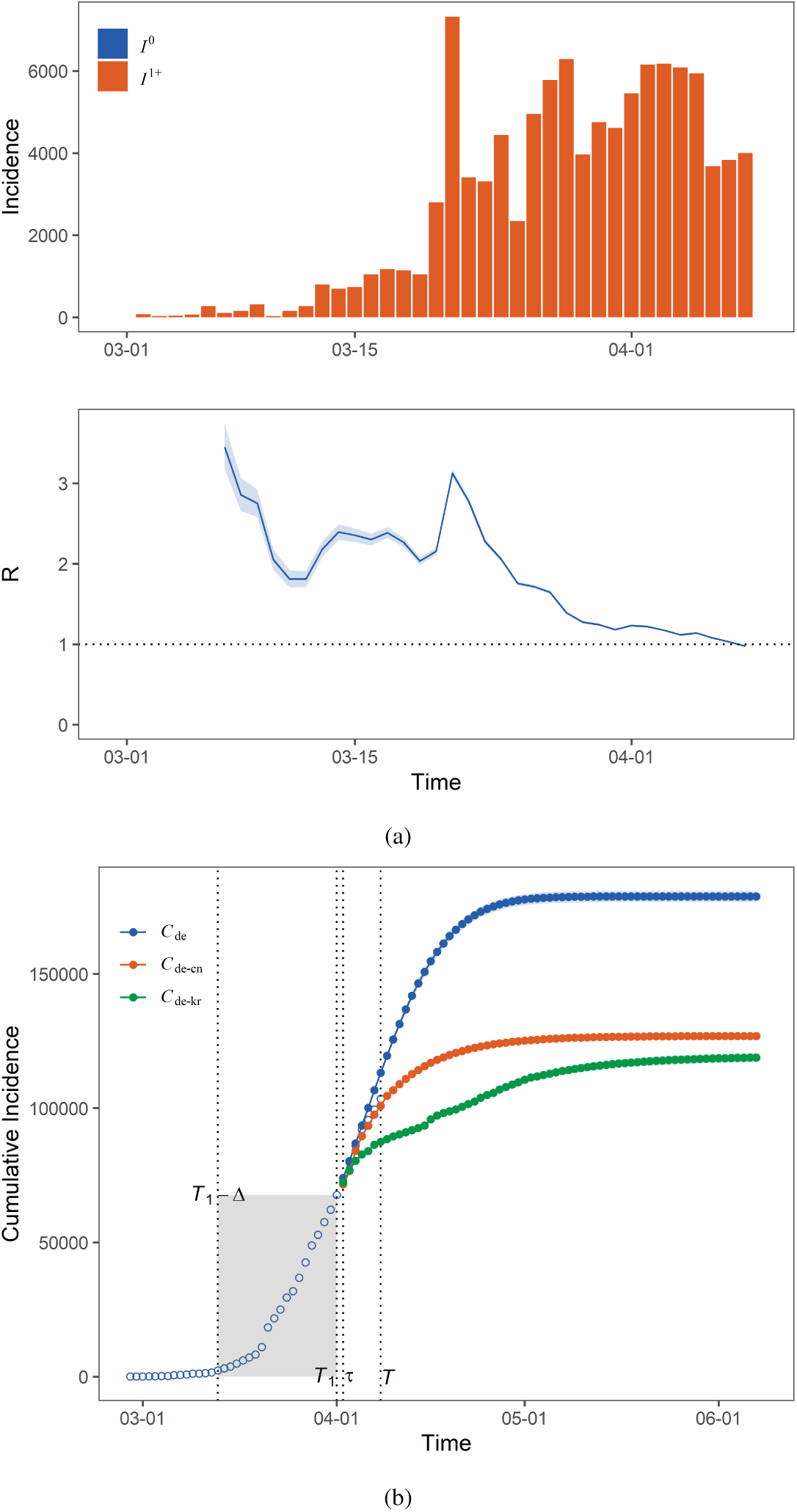
(a) Evolution of the number of cases and the time-varying reproduction number *R*(*t*) in Germany. (b) Accumulated case numbers at a time *T* and at the end of epidemics based on extrapolated *R_de_* (*t*) (blue line), *R_de−cn_* (*t*; *τ*) (orange line), and *R_de−kr_* (*t*; *τ*) (green line).

**Figure 5:**
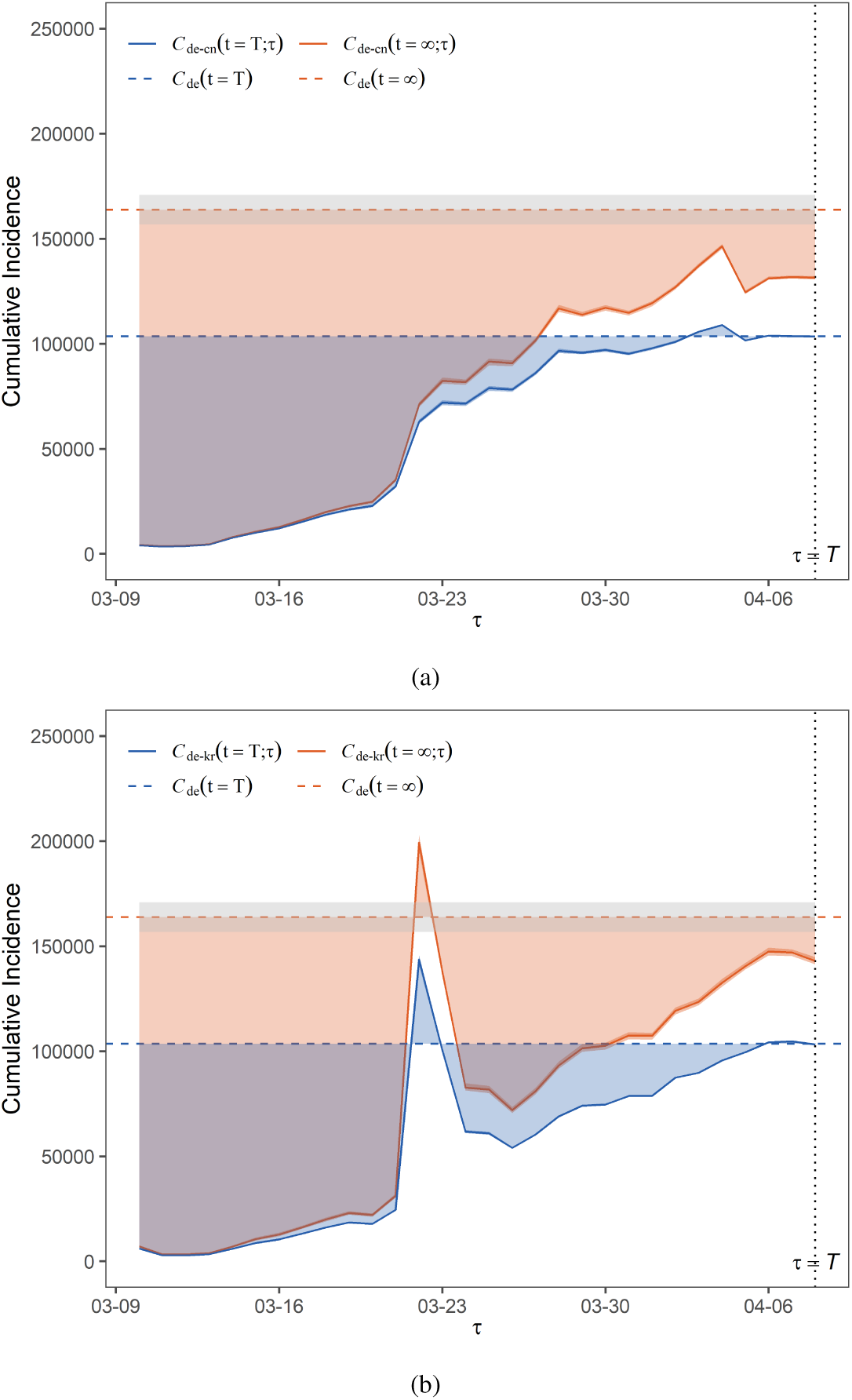
Evolution of the reduced infections in Germany with *τ* based on *R_de−cn_* (*t*; *τ*) (a) and *R_de−kr_* (*t*; *τ*) (b)

**Figure 6:**
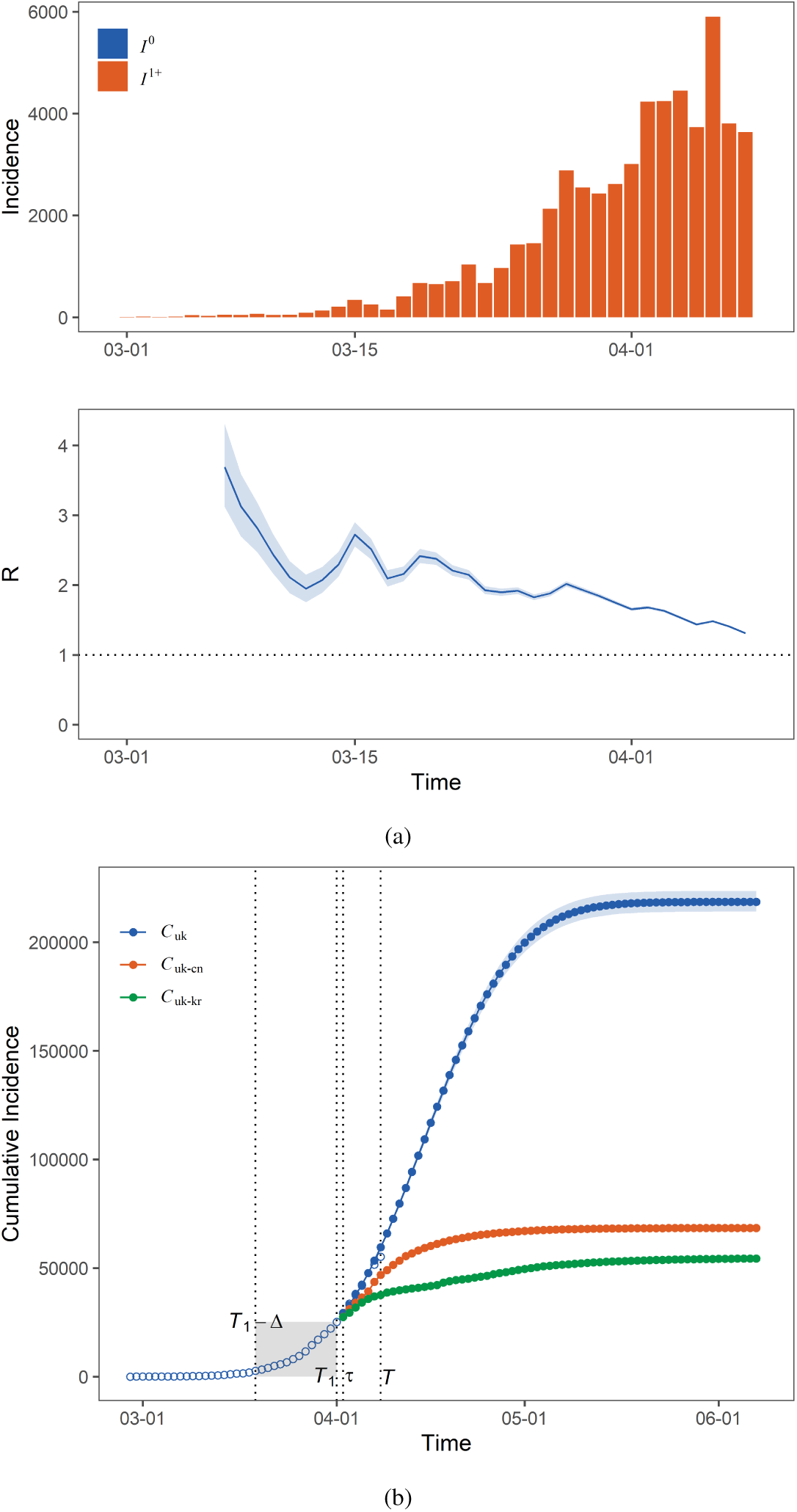
(a) Evolution of the number of cases and the time-varying reproduction number *R(t*) in UK (b) Accumulated case numbers at a time *T* and at the end of epidemics based on extrapolated *R_uk_* (*t*) (blue line), *R_uk−cn_* (*t*; *τ*) (orange line), and *R_uk−kr_* (*t*; *τ*) (green line).

**Figure 7:**
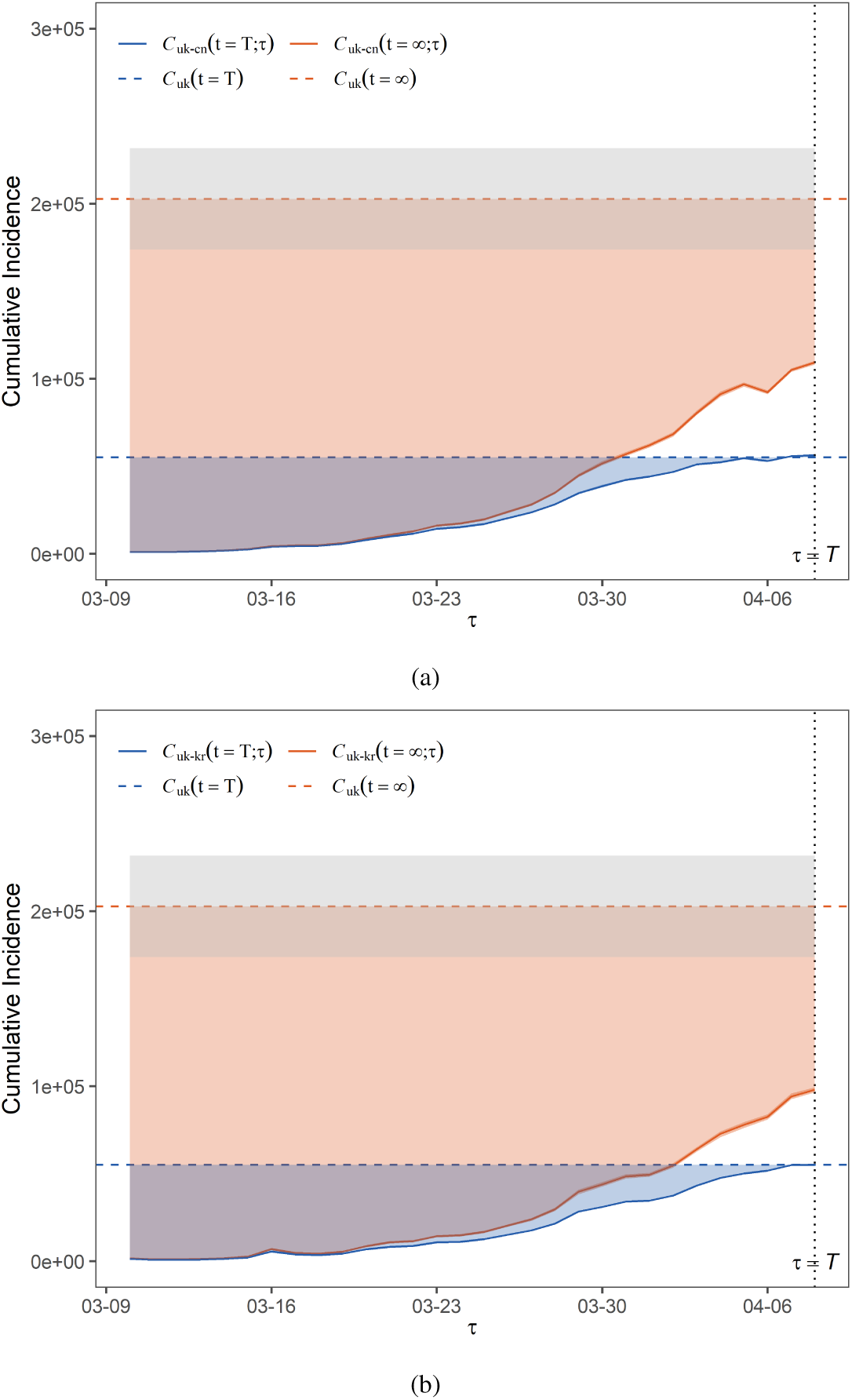
Evolution of the reduced infections in UK with *τ* based on *R_uk−cn_* (*t*; *τ*) (a) and *R_uk−kr_* (*t*; *τ*) (b)

**Figure 8:**
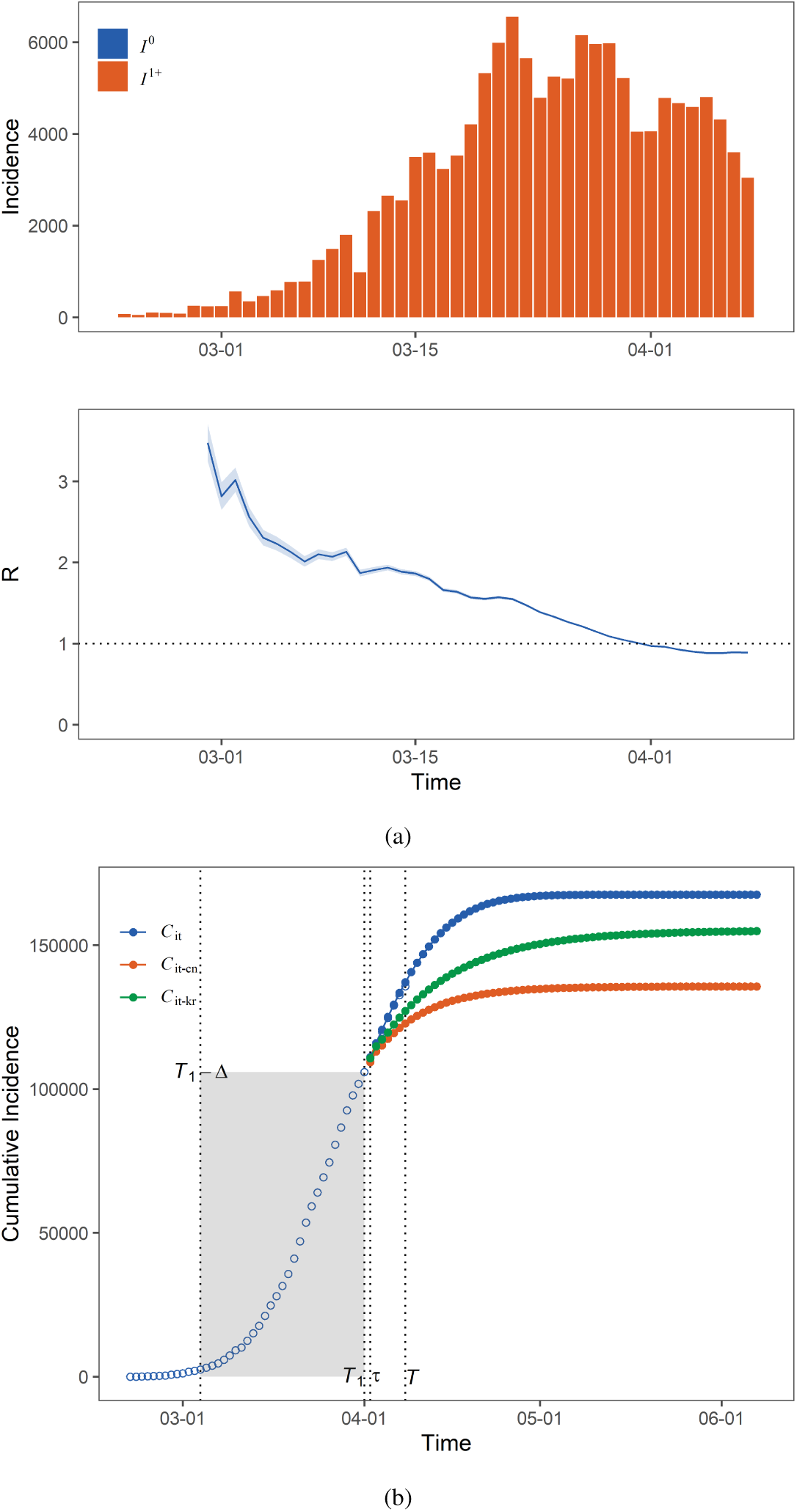
(a) Evolution of the number of cases and the time-varying reproduction number *R*(*t*) in Italy (b) Accumulated case numbers at a time *T* and at the end of epidemics based on extrapolated *R_it_* (*t*) (blue line), *R_it−cn_* (*t*; *τ*) (orange line), and *R_it−kr_* (*t*; *τ*) (green line).

**Figure 9:**
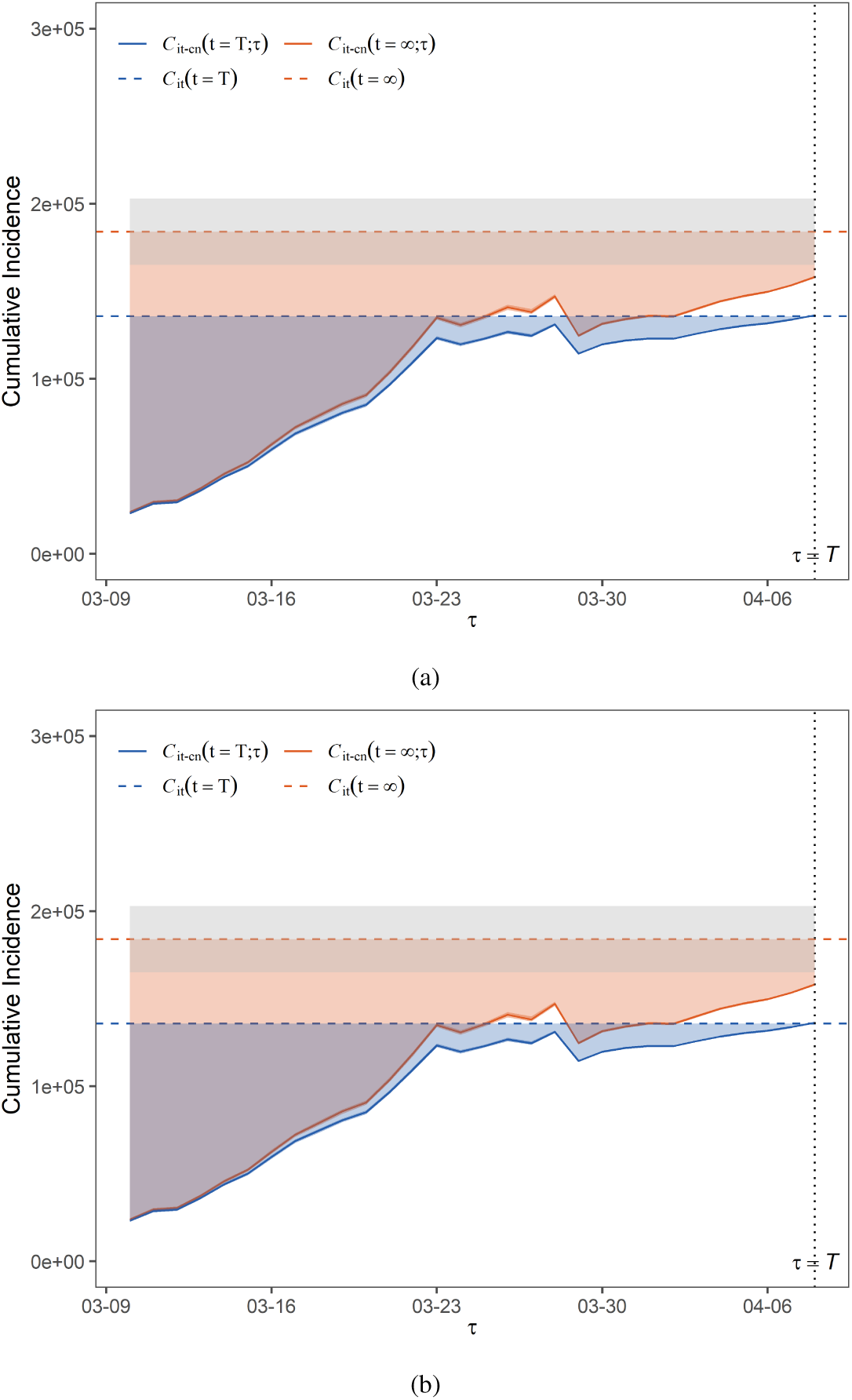
Evolution of the reduced infections in UK with *τ* based on *Rit−cn* (*t*; *τ*) (a) and *R_it−kr_* (*t*; *τ*) (b)

For *C_us_* (*t* = ∞), the final accumulated case numbers of the US shown in Fig. 3, we run an extrapolation for *R_us_ (t > T*) from the chosen data (*T* − ∆, *T*] for various ∆ and then plot the average value of accumulated case numbers (averaged over all values of ∆) and the standard deviation. Since we only need this *C_us_* (*t* = ∞) as a reference value instead of truly performing this prediction in this work, we are satisfied with this simple extrapolation.

**Connecting** *R_cn_* (*t*) **and** *R_us_* (*t*). When we connect *R_cn_* (*t*) to *R_us_* (*t*) at time *τ*, we first apply a *W*-day time window average of both time serials and then check whether there is a match of 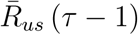 in the whole 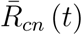. If there are such matched values, we take the last one, say, at *t*_0_, and connect the rest of *R_cn_* (*t* ≥ *t*_0_) to the right-hand side starting from *R_us_* (*τ*) so that we eventually arrive at *R_us−cn_* (*t*, *τ*) for all *t*.

If there is not yet such a matched value, which means that 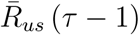 is larger than all values of 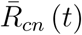, we first left-extrapolate 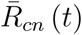 to a larger value to find a matched value and then connect them as described in the above paragraph.

The reason we perform the extrapolation and the connecting via *W*-day time window average time series is that the time series of daily values are considerably fluctuating and noisy.

